# Dynamic Contact Networks of Residents of an Urban Jail in the Era of SARS-CoV-2

**DOI:** 10.1101/2023.09.29.23296359

**Authors:** Samuel M. Jenness, Karina Wallrafen-Sam, Isaac Schneider, Shanika Kennedy, Matthew J. Akiyama, Anne C. Spaulding

## Abstract

**Background:** In custodial settings such as jails and prisons, infectious disease transmission is heightened by factors such as overcrowding and limited healthcare access. Specific features of social contact networks within these settings have not been sufficiently characterized, especially in the context of a large-scale respiratory infectious disease outbreak. The study aims to quantify contact network dynamics within the Fulton County Jail in Atlanta, Georgia, to improve our understanding respiratory disease spread to informs public health interventions.

**Methods:** As part of the Surveillance by Wastewater and Nasal Self-collection of Specimens (SWANSS) study, jail roster data were utilized to construct social contact networks. Rosters included resident details, cell locations, and demographic information. This analysis involved 6,702 residents over 140,901 person days. Network statistics, including degree, mixing, and turnover rates, were assessed across age groups, race/ethnicities, and jail floors. We compared outcomes for two distinct periods (January 2022 and April 2022) to understand potential responses in network structures during and after the SARS-CoV-2 Omicron variant peak.

**Results:** We found high cross-sectional network degree at both cell and block levels, indicative of substantial daily contacts. While mean degree increased with age, older residents exhibited lower degree during the Omicron peak, suggesting potential quarantine measures. Block-level networks demonstrated higher mean degrees than cell-level networks. Cumulative degree distributions for both levels increased from January to April, indicating heightened contacts after the outbreak. Assortative age mixing was strong, especially for residents aged 20–29. Dynamic network statistics illustrated increased degrees over time, emphasizing the potential for disease spread, albeit with a lower growth rate during the Omicron peak.

**Conclusions:** The contact networks within the Fulton County Jail presented ideal conditions for infectious disease transmission. Despite some reduction in network characteristics during the Omicron peak, the potential for disease spread remained high. Age-specific mixing patterns suggested unintentional age segregation, potentially limiting disease spread to older residents. The study underscores the need for ongoing monitoring of contact networks in carceral settings and provides valuable insights for epidemic modeling and intervention strategies, including quarantine, depopulation, and vaccination. This network analysis offers a foundation for understanding disease dynamics in carceral environments.

## INTRODUCTION

Custodial settings like jails, where arrested persons await trial, and prisons, which house sentenced persons, have long been associated with increased rates of infectious disease risk. Risk can be driven by both high baseline prevalence among those entering these systems and also high rates of contacts within them (1,2). For airborne infectious diseases, risks include the high density of persons within confined spaces, ongoing movement of persons progressing through the legal system (e.g., court visits), and frequent opportunities for new introductions of community-derived infections through correctional officers and visitors (3,4). The H1N1 pandemic resulted in correctional facility outbreaks driven by overcrowded living conditions and limited healthcare access for incarcerated individuals (5). For COVID-19, these features predictably created the context for numerous, large outbreaks (6,7). During the early pandemic period, 16 of the 20 largest COVID-19 clusters in the United States occurred in carceral facilities (8), including jails and prisons in Illinois, Ohio, California, and Texas (9,10).

Despite this increased environmental risk for SARS-CoV-2 and other respiratory infectious disease transmission in carceral settings, access to and use of prevention tools are suboptimal. It is often a challenge to isolate sick individuals and there is decreased access to cleaning supplies and other non-pharmaceutical interventions (NPI) (11). Another structural NPI approach has focused on reducing jail and prison population sizes (decarceration of existing populations) and arrests (reducing jail inflow). The combination have been effective at minimizing SARS-CoV-2 spread (12–14). While vaccines have been delivered among carceral populations, their availability was often delayed. For instance, in Georgia, correctional residents and officers were not highly prioritized in the initial distribution plan (15). After more widespread availability, hesitancy still was a barrier to uptake, including among prison staff (16,17). In California, for example, 61% of custodial staff and 36% of health care staff remained unvaccinated as of June 2021, even after widespread vaccine availability to the general public (18).

In spite of the potential role of infection control in custodial settings for disease risk, there are limited empirical data available to characterize the contact networks of potential exposures to SARS-CoV-2 and other respiratory pathogens (10). Much of the existing literature has described the overall rates of jail intake or discharge, summarized as an average daily contact rate (6,19). While those statistics are useful for broadly understanding transmission potential, they are insufficient to identify the epidemiological transmission dynamics that drive spread of disease *within* sectors of a jail or prison (20). Those dynamics depend on the patterns of contact networks, which may be characterized both in cross-sectional intensity (e.g., network degree), patterns of contacts between populations of the jail (e.g., age-related assortative mixing), and the rates of turnover of those contacts (e.g., average time spent within different jail sectors) (21). Network features may be additionally characterized at multiple layers (e.g., cells versus cell-blocks), with varying transmission potential based on the spatial density of contacts (22). Finally, it is important to investigate how contact network structures change over time, for example in response to large disease outbreaks (10). Describing these temporal network features will be critical to evaluate the potential impact of distancing-based approaches (isolation and quarantine) in jail and prison settings with suboptimal uptake of vaccines and other NPI.

In this study, we describe the contact networks of residents of the Fulton County Jail (FCJ) in Atlanta, Georgia between October 2021 and May 2022. The facility, with a rated capacity of 2600 in two-person occupancy cells, experienced historical overcrowding, averaging 104% during this period; this forced housing overflow populations in the open, common areas of housing units rather than cells. Despite ongoing efforts to house excess population in neighboring county jails, overcrowding could have increased the SARS-CoV-2 transmission potential. The lack of free space within FCJ also limited options for isolation and quarantine (23). Infected persons were often “sheltered in place,” kept in their cell with no new persons from their cell block assigned, and no new persons from other areas of the jail were assigned to their cell block, given the limited areas to isolate infected persons.

Our primary objective was to summarize the features of the contact networks with respect to three network statistics (degree, assortative mixing, and dissolution) overall, and then stratified by resident demographics, and jail layer (cell versus blocks). Our secondary objective was to compare these network features in January 2022, at the height of the Omicron SARS-CoV-2 wave in Atlanta, to a control time period in April 2022, when SARS-CoV-2 transmission had substantially declined (24). The broad goal of this study is to support public health interventions for respiratory infectious disease prevention within jails, by providing parameters relevant for epidemic modeling within these settings.

## METHODS

This analysis was part of a parent study, Surveillance by Wastewater and Nasal Self-collection of Specimens (SWANSS), the goal of which was to characterize the epidemiology of SARS-CoV-2 in the FCJ system using swab-based PCR testing and wastewater sampling. This analysis used data from October 2021 to May 2022, but also focused on two equally sized date spans to compare resident contact networks during and after the major SARS-CoV-2 Omicron variant transmission wave (25): 11 days in January 2022 and 11 days in April 2022. Contact networks are represented through jail roster data that define the primary residential location of incarcerated residents on any given day. These data are used to reconstruct the contact networks defined as shared physical space. This phase of the SWANSS project activities were approved by the Emory Institutional Review Board.

### Roster Data

FCJ provided study staff with jail roster data on a routine basis throughout the study period. While the primary purpose of these rosters was to identify the names and locations of jail residents to screen for SARS-CoV-2, we used these rosters to define spatial contact networks relevant for disease transmission. Daily rosters contained the names, unique identifier code, intake date, date of birth, sex, race/ethnicity, and cell location of each resident in FCJ. Cell location was coded hierarchically to identify the jail cell, cell block, floor, and tower. This analysis focused on the main FCJ building, which is comprised of two towers, each tower with 7 floors, and each tower-floor with 6 cell-blocks, which are housing units with two tiers of approximately 19 two-person cells. Residents within blocks could be assigned to an individual cell, usually with one other cell-mate. Alternatively, they could be assigned to the shared common area within the block if individual cells were unavailable. There were demographic and behavioral differences across the floors and blocks. Those having an aggressive institutional history, often the younger adults, or more serious criminal charges, were housed on the higher floors.

Our data flow diagram (**Figure 1**) shows the construction of our primary analytic datasets from the rosters. The combined dataset included 8,525 unique residents, contributing 160,157 person days over the study timeframe between October 2021 and May 2022. We excluded records from the analytic dataset for three reasons. First, data with missing location were excluded (2,938 person days); these were usually newly arrived jail residents at intake who were not yet assigned a housing location. Second, residents who were housed in special locations within the jail were excluded (15,675 person-days). These locations primarily were medical and psychiatric units within the FCJ where the contact networks were systematically different from the networks in the standard jail locations. Other special locations were holding cells for residents being transported to different facilities. Third, we excluded women from this analysis (643 person-days). A small number of women were housed within the main FCJ building, primarily for medical reasons, with most incarcerated women were housed in an annex building not considered here. That left a total of 6,702 unique residents, contributing 140,901 person days.

**Figure 1.**
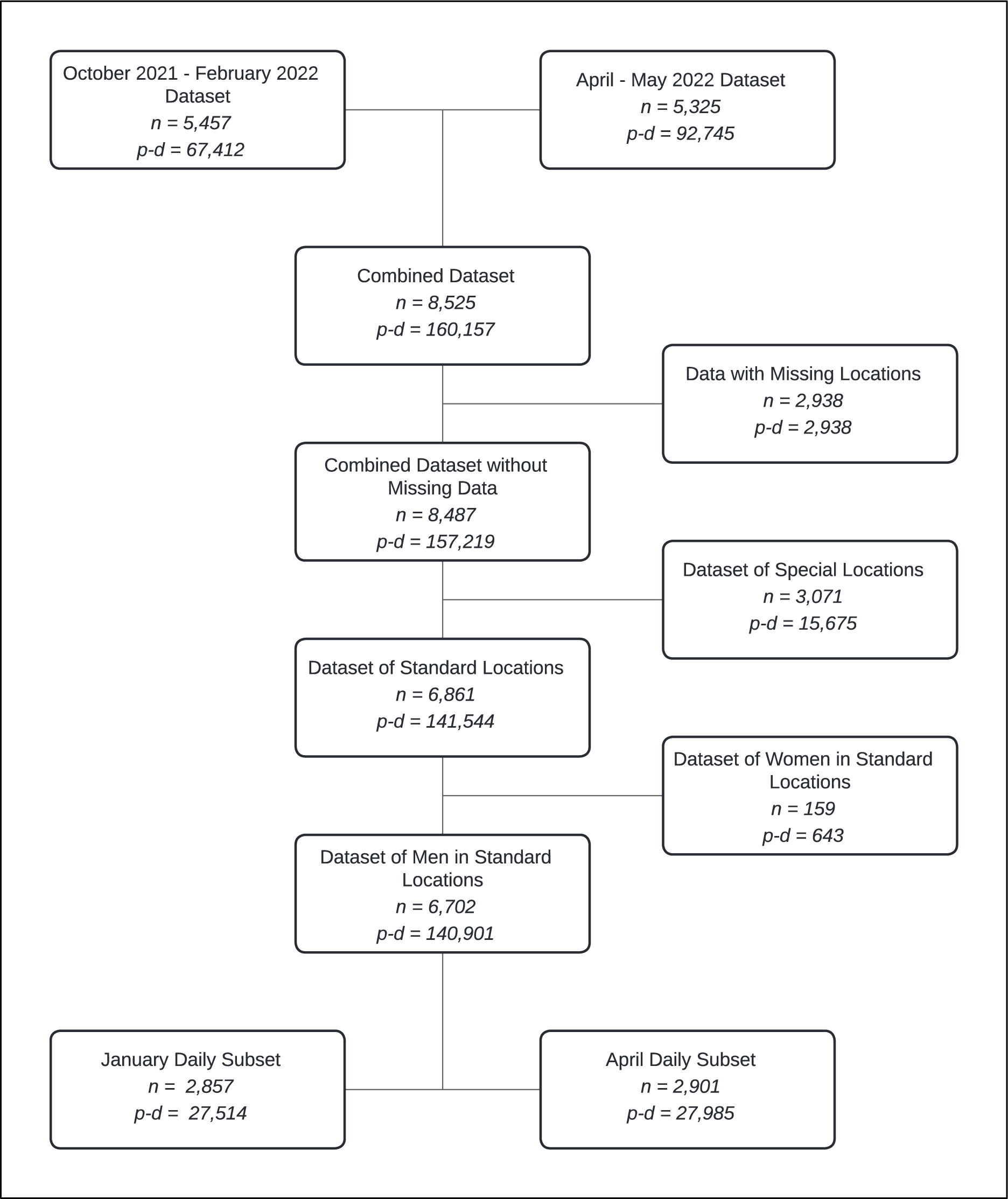
PRISMA diagram of Fulton County Jail roster data used in analysis, by overall unique persons (n) and person days (p-d). From two starting datasets spanning October 2021 to May 2022, the combined dataset included 8,525 unique jail residents, contributing 160,157 days. After exclusion of missing and non-standard locations and limited records to men, there were 6,702 unique persons. For Omicron wave comparisons, we further stratified the rosters into January and April subsets.

### Network Measures

We defined contact networks as those involving shared physical areas of the FCJ building on any day and over time. This was based on the understanding that SARS-CoV-2 transmission requires sustained physical proximity (26). Here we focused on cell-level and block-level contact networks; contacts between residents across blocks on any day were uncommon. A schematic of the FCJ contact networks in this analysis is presented in **Figure 2**. Residents are represented as nodes, and contacts are represented as edges between nodes. The node set is the same across the block-level and cell-level networks, but the edge set for the block-level network is a superset of the edge set for the cell-level network (all cell mates were block mates but not vice versa). Residential location could change daily, as residents were moved to different areas of the building. We identified a movement in location as the same resident listed on sequential rosters in a different cell. A resident could therefore have a different contact network daily. If rosters were missing on any intermediate day (interval censoring), we assumed stability of the residential location in the unobserved interval between rosters.

**Figure 2.**
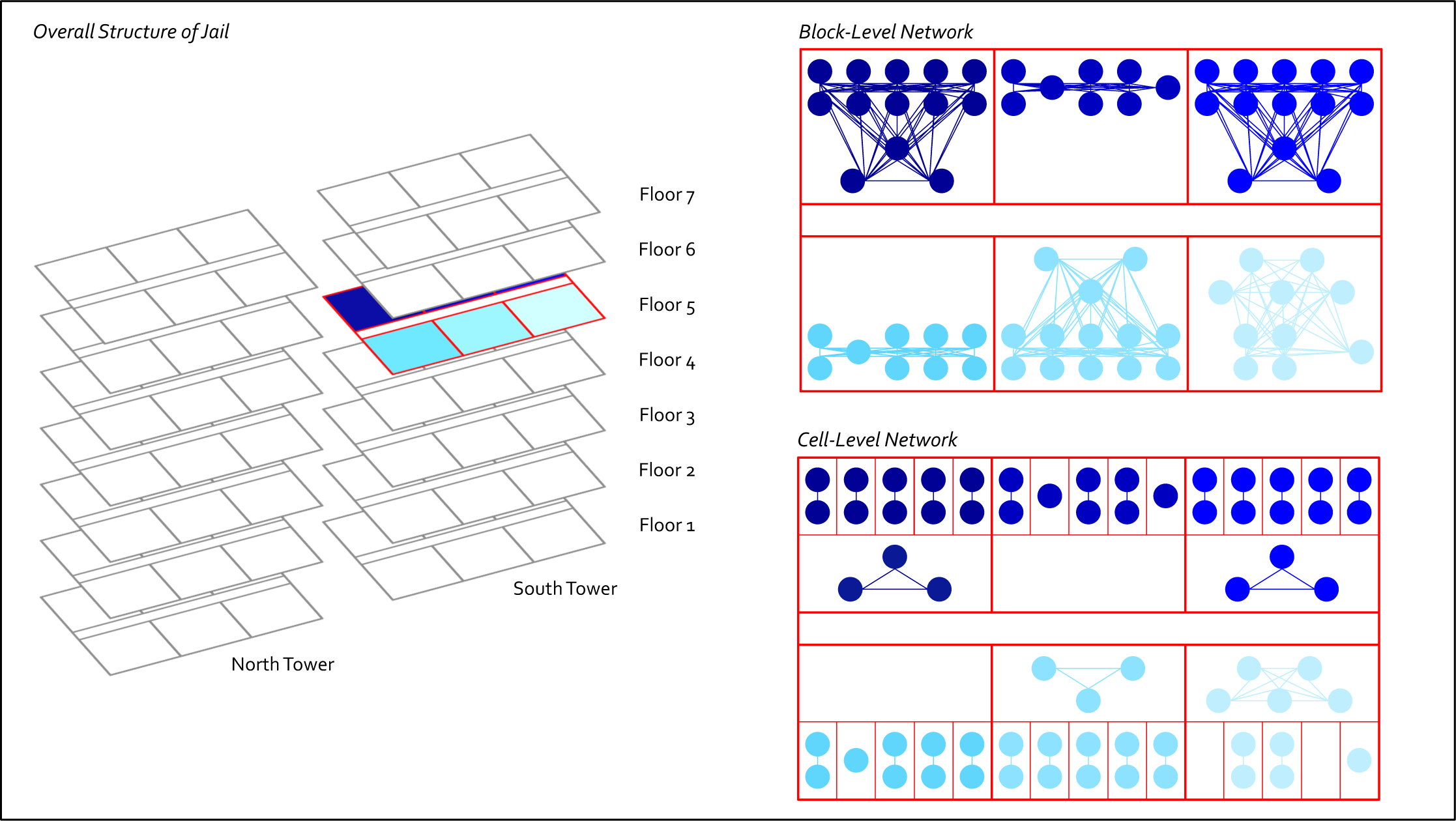
Schematic of Fulton County Jail (FCJ) representation in analysis. The FCJ main building are structured into two towers (North and South), each of which contain 7 floors. Each floor contains up to 6 blocks, and each (standard) block contains up to 19 cells. Transmission-relevant social contacts occur among residents sharing a cell, but can also occur among residents in the same block during free time spent outside the cell. We visualize the block-level and cell-level networks accordingly.

We quantified three common dynamic network statistics: *degree*, *mixing*, and *dissolution*. All three measures were stratified by age group, race/ethnicity, and floor number. Our primary degree measure, momentary active degree, was the cross-sectional count of unique contacts per person on any given date within either the cell or block layer. Degree was also calculated over the January and April date spans (dynamic degree) to count the number of unique contacts over each 11-day span. Mixing was evaluated with respect to age, as this was the demographic most relevant for SARS-CoV-2 acquisition and severe COVID-19 disease (27). We categorized residents into decades of age, based on date of birth at their initial roster. Then we divided the total contacts of each age group into decades of age for their contacts. To do this, we organized age of residents into a symmetrical undirected mixing matrix. Finally, network dissolution rates quantified the turnover of cell-level, block-level, and jail-level contacts per day.

Dissolution rates were calculated as the reciprocal of the average time spent in the location for any given “time spell.” For instance, if a resident was located in two cells across the observed time—one spell for 10 days and the other for 20 days—the average duration across spells was 15 days and thus, the average rate of dissolution was 0.067 per day. We further categorized residents as short-term or long-term, where the latter indicates having a single spell of activity censored on both sides of available rosters.

### Statistical Analyses

This is a descriptive statistical analysis, intended to provide empirical estimates of the three network statistics, quantify how they differed across demographics, and evaluate how they changed over time. The rosters are a census of the FCJ resident population on any day, so there is no inferential uncertainty with respect to sampling. For network degree, we estimated the mean of the degree distribution across residents on any given date. We averaged degree across the entire study time frame and across the two 11-day spans in January and April to understand whether contact patterns changed over time. For age mixing, we generated a standard age mixing matrix, which calculates the relative proportion of contacts between each age group pairing. Numerically, we quantified the relative size of the diagonal of this mixing matrix, which is the fraction of contacts that occurred within the same age group (age assortative mixing). We show how assortative mixing varied across the two time spans to understand age segregation during these different periods. Finally, dissolution was quantified as the average count of movements per resident per day. Dissolution rates are subject to both interval censoring (in the case that unobserved changes between available rosters occurred) and boundary (left and right) censoring (in the case that future changes before or after the final roster occurred).

## RESULTS

The descriptive characteristics of the jail residents are provided in **Table 1**. Across the full study time span (October 2021 to May 2022), a total of 6,702 unique residents were identified in the rosters. Nearly two-thirds of residents were between the ages of 20 and 39 years old (63.4%). Residents overwhelmingly identified as Black race (88.5%). Across the floors of the FCJ building, most had been on Floor 2, which is used as a temporary placement floor before longer-term cell reassignment. The unique population sizes were similar for the two 11-day spans in January and April 2022. The age and race distributions were also similar.

**Table 1.** Descriptive Characteristics of Fulton County Jail Residents.

| Characteristic | January 2022 Span<br>*n* (%) | April 2022 Span<br>*n* (%) | October 2021–<br>May 2022<br>*n* (%) |
| --- | --- | --- | --- |
| **Sex** |  |  |  |
| Male | 2857 (100%) | 2901 (100%) | 6702 (100%) |
| **Age<sup>1</sup>** |  |  |  |
| 17–20 | 191 (6.7%) | 186 (6.4%) | 396 (5.9%) |
| 20–29 | 981 (34.3%) | 979 (33.7%) | 2145 (32.0%) |
| 30–39 | 860 (30.1%) | 891 (30.7%) | 2103 (31.4%) |
| 40–49 | 459 (16.1%) | 463 (16.0%) | 1084 (16.2%) |
| 50–59 | 264 (9.2%) | 262 (9.0%) | 697 (10.4%) |
| 60–84 | 102 (3.6%) | 120 (4.1%) | 277 (4.1%) |
| **Race<sup>2</sup>** |  |  |  |
| Black | 2551 (89.3%) | 2603 (89.7%) | 5930 (88.5%) |
| Other | 26 (0.9%) | 25 (0.9%) | 62 (0.9%) |
| White | 280 (9.8%) | 273 (9.4%) | 710 (10.6%) |
| **Floor<sup>3</sup>** |  |  |  |
| Ever on Floor 1 | 278 (9.7%) | 251 (8.7%) | 955 (14.2%) |
| Ever on Floor 2 | 696 (24.4%) | 744 (25.6%) | 3412 (50.9%) |
| Ever on Floor 3 | 425 (14.9%) | 458 (15.8%) | 1258 (18.8%) |
| Ever on Floor 4 | 458 (16.0%) | 458 (15.8%) | 1328 (19.8%) |
| Ever on Floor 5 | 456 (16.0%) | 467 (16.1%) | 1096 (16.4%) |
| Ever on Floor 6 | 429 (15.0%) | 432 (14.9%) | 724 (10.8%) |
| Ever on Floor 7 | 297 (10.4%) | 312 (10.8%) | 601 (9.0%) |
| **Duration<sup>4</sup>** |  |  |  |
| Long-Term | – | – | 1024 (15.3%) |
| Short-Term | – | – | 2877 (42.9%) |

<sup>1</sup> A person's age is calculated as of the first day they appear in a roster during the timeframe of interest
<sup>2</sup> The 'Other' race category includes people whose race was recorded as Asian, Hispanic, Middle Eastern, Native Hawaiian or Other Pacific Islander, Indian, Multiracial, Other, or Unavailable
<sup>3</sup> People are defined as having been 'Ever on Floor *n*' if they were on Floor *n* for at least one day during the timeframe of interest. Thus, these categories are not mutually exclusive.
<sup>4</sup> People are considered 'long-term' if they have a single spell of activity that is censored on both sides (i.e., if they are in every roster); they are considered short-term if they have at least one spell of activity that is uncensored on both sides. (Some people – i.e., those who have only a left-censored spell and/or a right-censored spell – do not fall into either category.)

**Table 2** and **Figure 3** provide information on the mean degree across the full time span and the two 11-day spans. The overall mean degree was 1.95 in the cell-level networks, meaning on average that residents had nearly 2 cell-level contacts on any given day. Across time, mean degree was higher among older residents, from 1.26 among residents 20 years of age and younger to 2.74 for residents 60 years of age or older. Comparing time spans, the cell-level degree was 20% higher in the April span (mean degree = 2.07) than in the January span (mean degree = 1.72). Degree was relatively stable within each span (**Figure 3**). The rate of temporal changes in mean degree was greater for older age residents: mean degree increased 41% from January to April among residents aged 60 or older, compared to increasing only 18% among residents 20 or younger. Cell-level mean degree was comparable for residents of Black race, but was lower for non-White, non-Black “other race” residents (who comprised <1% of the population). Across floors, mean degree was generally less on the higher floors, and the least on Floor 7. Mean degree at the block-level network was considerably higher than the cell-level network. On average, there were 34.11 contacts on any day in the block-level network. For this network, there were only minor increases (4%) in mean degree between January and April. Compared to the cell-level networks, the block level networks also did not show strong gradients among older residents, or changes over time for older residents.

**Figure 3.**
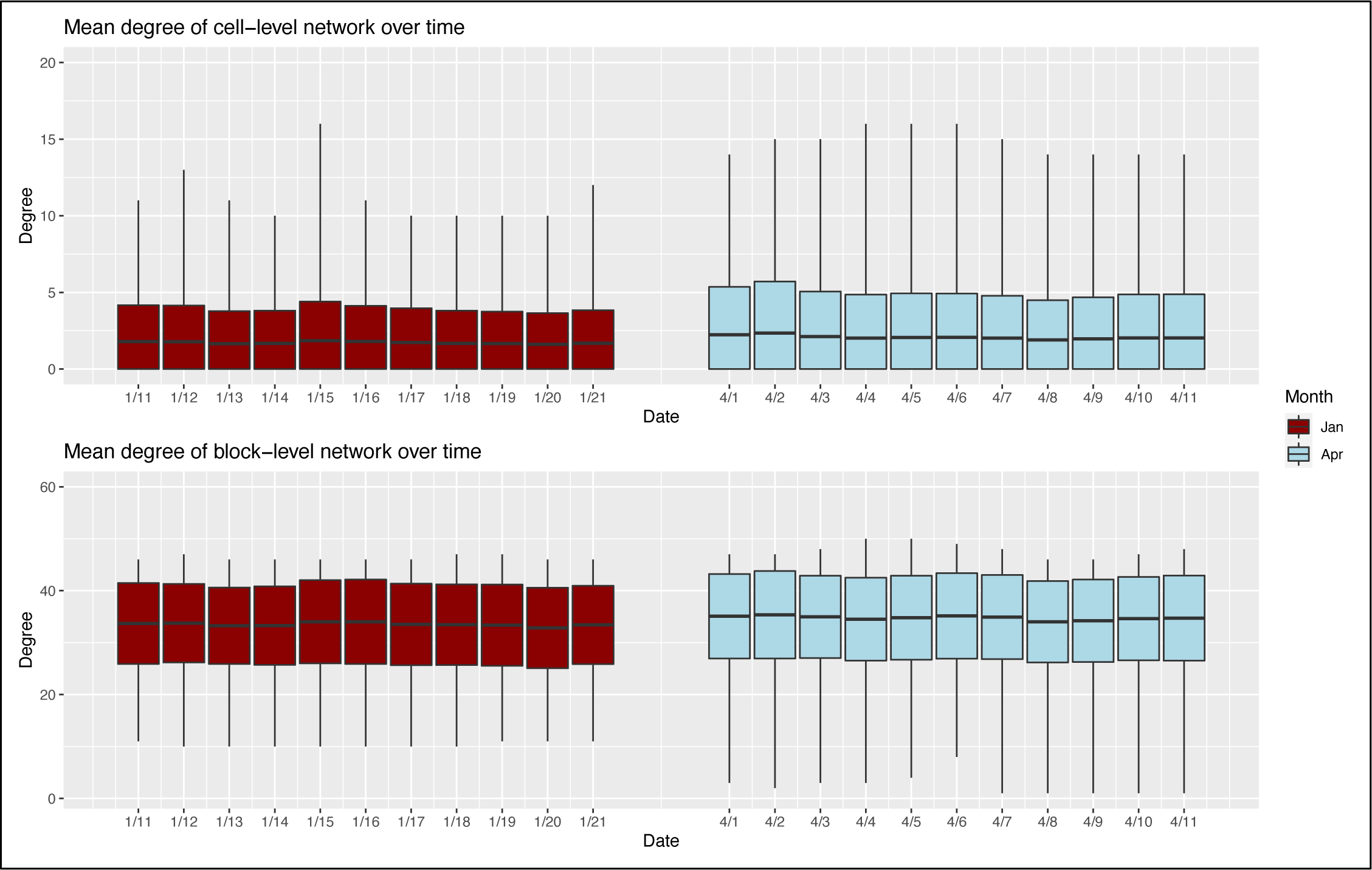
Social network degree distribution for cell-level and block-level networks during January and April roster dates. Boxes display the variation in mean degree over each date within January versus April.

**Table 2.** Mean Degree of Cell- and Block-Level Networks for Various Timeframes.

|  | Mean Degree of Cell-Level Network |  |  | Mean Degree of Block-Level Network |  |  |
| --- | --- | --- | --- | --- | --- | --- |
|  | January 2022<br>Span | April 2022<br>Span | October 2021–<br>May 2022 | January 2022<br>Span | April 2022<br>Span | October 2021–<br>May 2022 |
|  | *Mean (SD)<sup>1</sup>* | *Mean (SD)<sup>1</sup>* | *Mean (SD)<sup>1</sup>* | *Mean (SD)<sup>1</sup>* | *Mean (SD)<sup>1</sup>* | *Mean (SD)<sup>1</sup>* |
| **Overall** |  |  |  |  |  |  |
| Overall | 1.72 (2.23) | 2.07 (2.89) | 1.95 (2.69) | 33.50 (7.74) | 34.76 (8.08) | 34.11 (8.00) |
| **By Age** |  |  |  |  |  |  |
| 17–20 | 1.14 (1.53) | 1.34 (2.07) | 1.26 (1.84) | 28.63 (8.51) | 26.77 (9.44) | 27.72 (9.13) |
| 20–29 | 1.46 (1.91) | 1.79 (2.60) | 1.66 (2.32) | 32.04 (7.61) | 33.01 (8.47) | 32.63 (7.97) |
| 30–39 | 1.87 (2.39) | 2.13 (2.83) | 2.05 (2.74) | 34.66 (7.35) | 36.17 (7.02) | 35.35 (7.28) |
| 40–49 | 1.92 (2.34) | 2.11 (2.87) | 2.10 (2.78) | 35.36 (7.09) | 37.30 (6.27) | 36.48 (6.79) |
| 50–59 | 2.18 (2.64) | 3.10 (3.82) | 2.72 (3.40) | 35.43 (6.98) | 37.01 (6.82) | 36.42 (7.16) |
| 60–84 | 1.97 (2.63) | 2.78 (3.58) | 2.74 (3.47) | 33.86 (8.70) | 37.05 (7.01) | 35.33 (7.70) |
| **By Race** |  |  |  |  |  |  |
| Black | 1.74 (2.25) | 2.06 (2.87) | 1.96 (2.69) | 33.32 (7.79) | 34.57 (8.18) | 33.94 (8.07) |
| Other | 0.84 (0.37) | 1.63 (2.36) | 1.39 (1.64) | 34.91 (6.79) | 36.70 (5.62) | 36.10 (6.10) |
| White | 1.62 (2.10) | 2.21 (3.06) | 1.96 (2.72) | 35.06 (7.17) | 36.41 (6.96) | 35.65 (7.21) |
| **By Floor** |  |  |  |  |  |  |
| Floor 1 | 1.98 (2.41) | 1.98 (2.28) | 2.19 (2.58) | 35.77 (5.55) | 37.86 (4.43) | 37.14 (4.16) |
| Floor 2 | 1.98 (2.52) | 2.44 (3.06) | 2.28 (3.02) | 35.54 (6.63) | 37.27 (6.00) | 35.80 (6.60) |
| Floor 3 | 1.32 (2.12) | 2.61 (4.41) | 1.89 (3.15) | 28.26 (7.80) | 32.77 (9.94) | 30.32 (8.60) |
| Floor 4 | 1.59 (1.84) | 1.92 (2.41) | 1.89 (2.41) | 34.95 (5.97) | 35.47 (7.73) | 35.66 (7.22) |
| Floor 5 | 1.84 (2.20) | 2.09 (2.52) | 2.03 (2.51) | 35.32 (6.29) | 36.07 (7.79) | 35.69 (7.00) |
| Floor 6 | 2.00 (2.62) | 1.95 (2.50) | 1.99 (2.60) | 36.57 (7.63) | 36.27 (7.38) | 36.33 (7.69) |
| Floor 7 | 1.20 (1.03) | 1.21 (0.94) | 1.22 (1.00) | 25.52 (6.41) | 26.59 (5.00) | 26.29 (6.13) |

<sup>1</sup> The mean and the standard deviation of the degree distribution were calculated at each time point within the timeframe of interest for which a roster is available. These statistics were then averaged across the timeframe of interest. Thus, the number in parenthesis is the mean of the standard deviation at each time point (not the standard deviation of the mean at each time point).

**Figure 4** shows the cumulative degree distribution for the cell-level and block-level networks for the January and April spans. The distribution was higher overall for April compared to January for both cell-level and block-level networks. The increases in mean degree over time (**Table 2**) in both network layers occurred across the entire span of the degree distribution, not only to high-degree residents.

**Figure 4.**
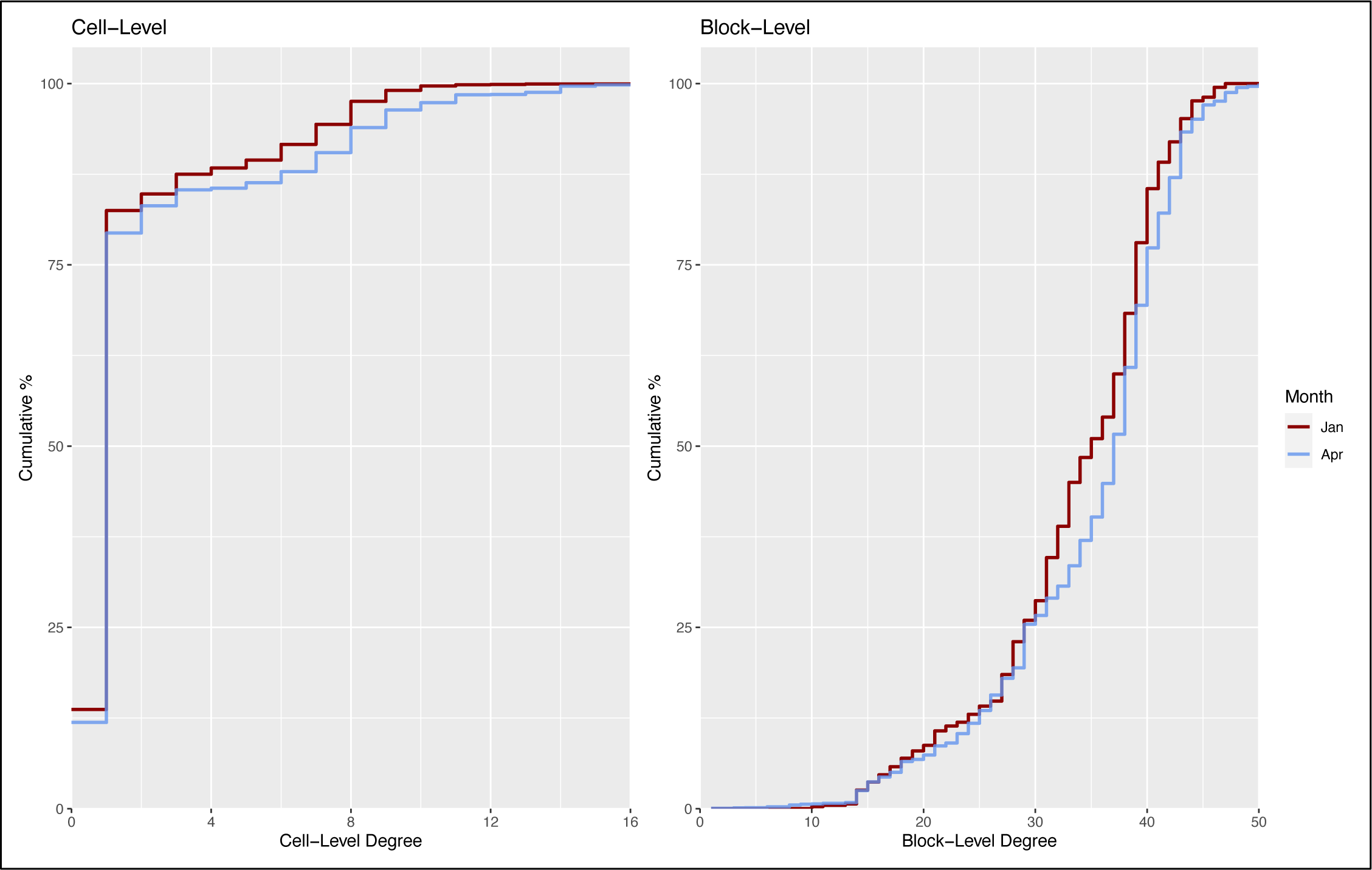
Cumulative degree distributions (proportion of residents with X or higher value of network degree), stratified by cell-level versus block-level degree definitions and January versus April 2022 date spans. The cumulative degree curves were higher at both cell-levels and block-levels for April compared to January.

**Figure 5** illustrates the dynamic network degree estimates for the January and April spans, considering the turnover in both cell-level and block-level networks over the 11 days in each span. The degree value on Day 1 in each panel is equivalent to cross-sectional mean degree values described above. Dynamic degree then increased over time due to turnover in the cell and blocks, as residents form new contacts in their new residential locations. The cell-level degree reached 2.64 in January and 3.50 in April, so the relative 11-day growth rate in dynamic degree was higher for April compared to January (69% versus 53%). For the block-level network, the degree was also higher over time for April than January, reaching 45.2 and 41.7, respectively on Day 11. Here again, the 11-day growth rate of the block level dynamic network was higher for April than January (30% versus 24%).

**Figure 5.**
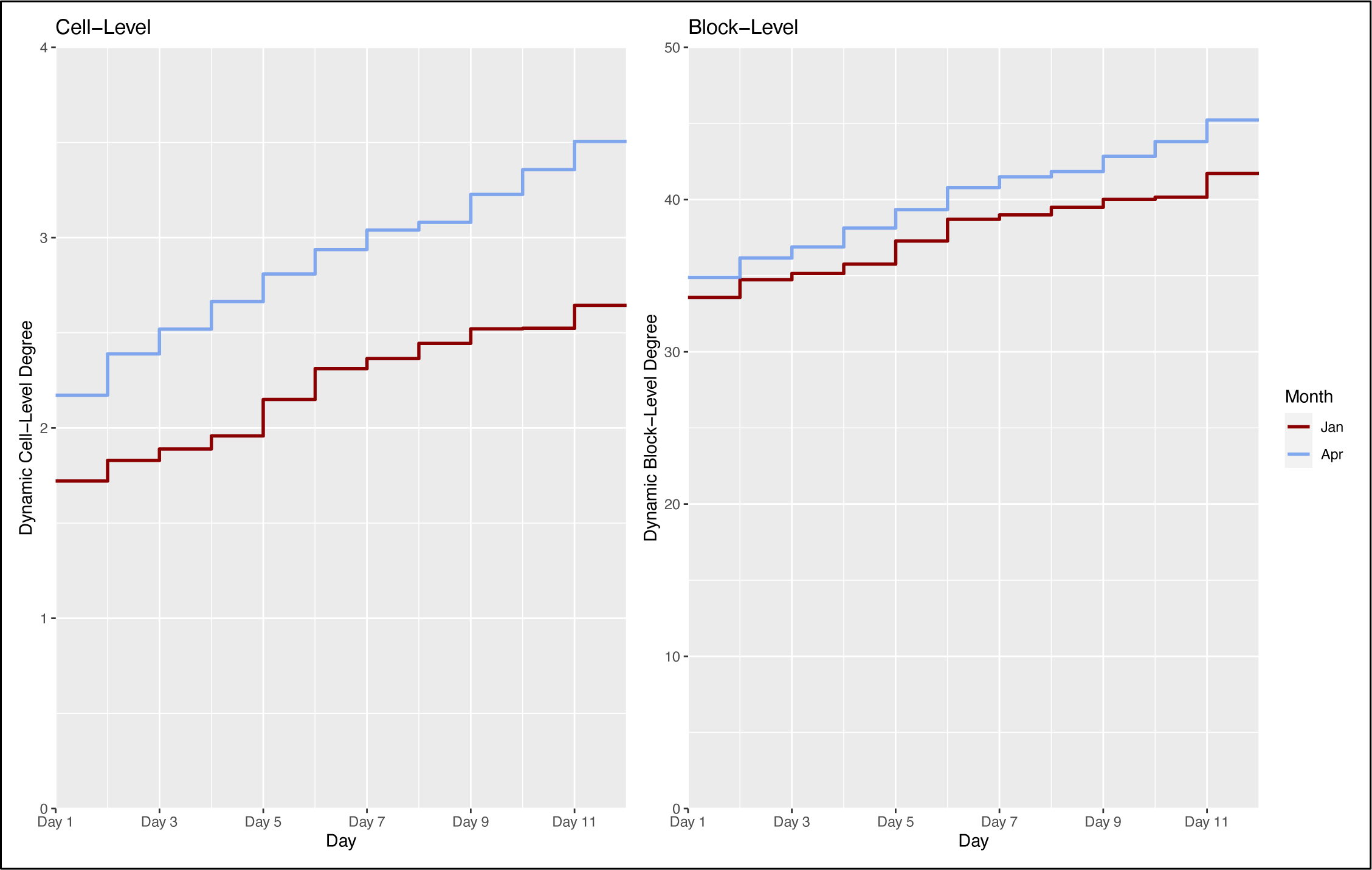
Mean dynamic network degree, counting the average number of unique contacts over the 11-day spans in January and April 2022 date ranges. Cumulative network degree starts at the cross-sectional value for cell-level and block-level degree on day 1, then monotonically increase as residents acquire new contacts due to movement within the jail.

We observed strong assortative age mixing across time spans and network layers. Figure 6 illustrates that a high proportion of network contacts occurred along the diagonals of the mixing matrices, indicating edges within similar age groups, although the line tapers downward at older age groups, indicating more contacts with middle-aged residents. Numerically, **Table 3** shows the fraction of network contacts that occurred within the same age group. Consistent with Figure 6, assortative mixing was strong, with 32% of both cell-level edges and block-level edges occurring within age group. Cell-level assortative mixing was strongest among middle-aged residents (aged 20 to 39), and weaker among the youngest and oldest residents. Cell-level assortative mixing was weaker among White race residents and was less for Floors 2 and 3. These demographic and floor patterns generally held for block-level edges. For both cell-level and block-level edges, the assortative mixing was slightly stronger for the January span than the April span, and this was driven by the highest levels of assortative mixing by residents aged 20–29.

**Figure 6.**
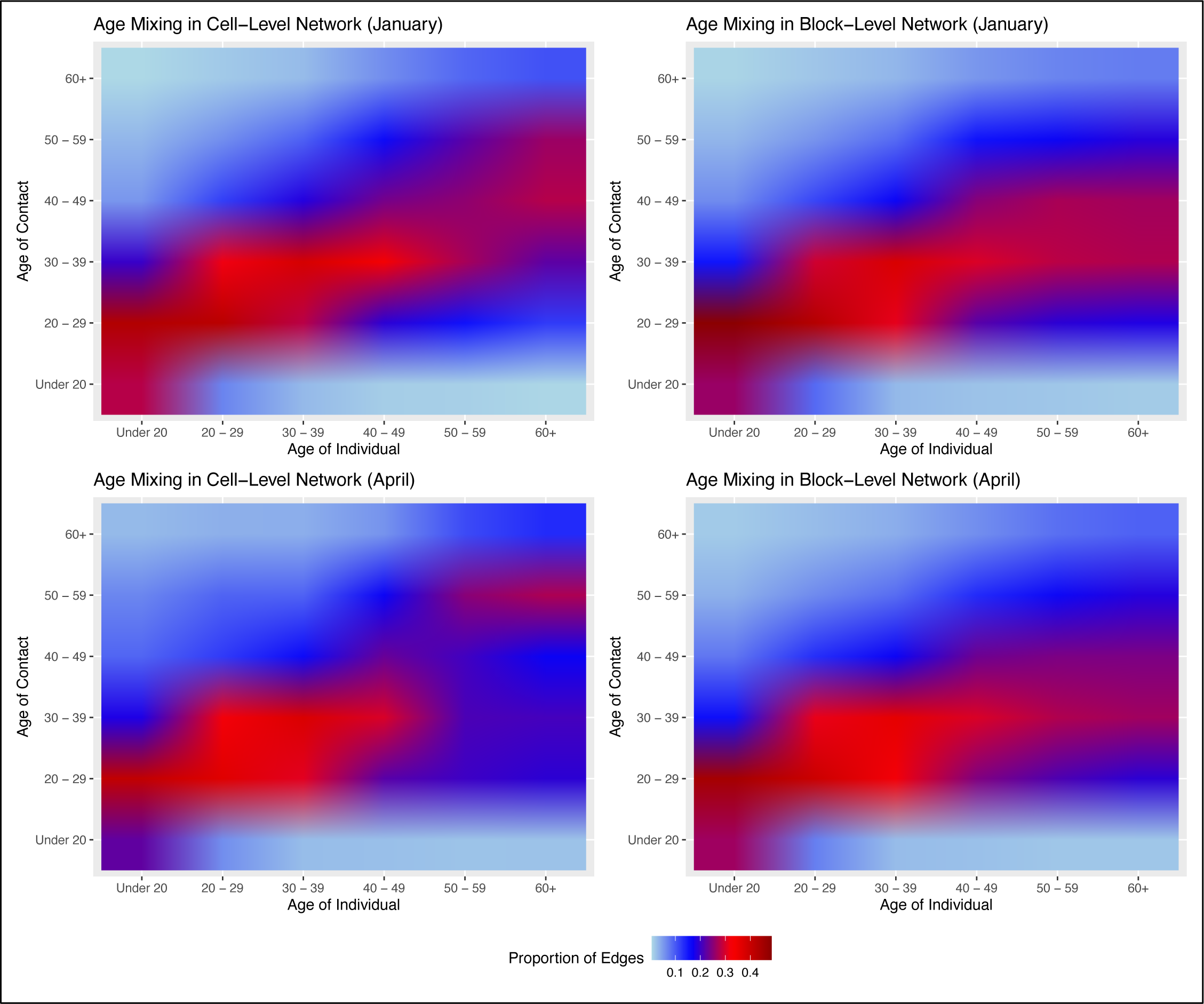
Age mixing in cell-level and block-level networks in the January and April roster date spans. Matrices are standardized to the total contacts in each age group, summing vertically to 1. There was stronger within-group age mixing in the cell-level compared to the block-level networks, and in both network types in January compared to April.

**Table 3.** Percent of Cell- and Block-Level Network Contacts within Same Age Group.

|  | Cell-Level Edges<sup>1</sup> |  |  | Block-Level Edges<sup>1</sup> |  |  |
| --- | --- | --- | --- | --- | --- | --- |
|  | January 2022<br>Span | April 2022<br>Span | October 2021–<br>May 2022 | January 2022<br>Span | April 2022<br>Span | October 2021–<br>May 2022 |
|  | % | % | % | % | % | % |
| **Overall** |  |  |  |  |  |  |
| Overall | 33.3% | 31.2% | 31.8% | 33.4% | 31.4% | 32.1% |
| **By Age** |  |  |  |  |  |  |
| 17–20 | 28.0% | 22.5% | 23.8% | 26.1% | 26.5% | 26.5% |
| 20–29 | 41.9% | 36.7% | 38.7% | 42.7% | 39.5% | 40.7% |
| 30–39 | 38.3% | 37.5% | 37.1% | 37.6% | 35.9% | 36.2% |
| 40–49 | 23.9% | 23.0% | 23.7% | 25.0% | 23.5% | 24.0% |
| 50–59 | 22.0% | 25.1% | 23.7% | 16.7% | 16.5% | 17.3% |
| 60–84 | 10.6% | 13.5% | 13.1% | 7.4% | 9.4% | 8.2% |
| **By Race** |  |  |  |  |  |  |
| Black | 33.8% | 31.9% | 32.4% | 34.0% | 32.1% | 32.8% |
| Other | 47.7% | 44.3% | 39.8% | 27.6% | 29.8% | 28.0% |
| White | 26.9% | 24.4% | 25.0% | 28.1% | 25.3% | 26.2% |
| **By Floor** |  |  |  |  |  |  |
| Floor 1 | 32.4% | 32.0% | 31.4% | 30.6% | 26.3% | 28.5% |
| Floor 2 | 32.1% | 29.9% | 28.8% | 27.7% | 26.1% | 26.6% |
| Floor 3 | 24.9% | 22.7% | 24.3% | 22.1% | 22.7% | 22.3% |
| Floor 4 | 34.0% | 30.6% | 33.3% | 38.7% | 34.8% | 35.7% |
| Floor 5 | 34.9% | 36.8% | 34.6% | 36.1% | 35.2% | 35.8% |
| Floor 6 | 36.4% | 35.7% | 36.0% | 39.2% | 39.2% | 38.7% |
| Floor 7 | 36.7% | 37.6% | 37.5% | 35.4% | 33.3% | 34.2% |

<sup>1</sup> These calculations are based off of the doubled edgelist (in which every edge is listed twice, e.g., once from node A to node B and once from node B to node A). The denominator is the number of these (doubled) edges for which the head falls into the given category; the numerator is the number of those edges for which the head and tail are in the same age group.

**Table 4** shows the rate of contact network dissolution at the cell and block levels, as well as for the jail overall, for the January and April spans. The rates of cell turnover were higher than the rates of block-level turnover, and the turnover rates within each level were higher in April than in January. The rates of releases from the jail were similar or slightly higher than to the rates of block-level turnover. The overall rates in April corresponded to a mean duration in cell before turnover (with an assumption of a geometric statistical distribution of time to events) of 38 days, compared to a mean block duration of 66 days, and a mean jail duration of 71 days. At the cell level, rates were slightly higher among younger residents than older residents, but similar across age. Rates were greatest among the lower floors at both the cell-level and block-level. In January the rate of release from jail were higher than the rate of block-level changes, implying there was more turnover in the jail overall than between blocks.

**Table 4.** Rates of Cell-Level, Block-Level, and Jail-Level Network Dissolution per Day.

|  | Rate of Cell-Level Changes |  | Rate of Block-Level Changes |  | Rate of Releases from Jail |  |
| --- | --- | --- | --- | --- | --- | --- |
|  | January 2022 | April 2022 | January 2022 | April 2022 | January 2022 | April 2022 |
|  | *Rate Per Day<sup>1</sup>* | *Rate Per Day<sup>1</sup>* | *Rate Per Day<sup>2</sup>* | *Rate Per Day<sup>2</sup>* | *Rate Per Day<sup>3</sup>* | *Rate Per Day<sup>3</sup>* |
| **Overall** |  |  |  |  |  |  |
| Overall | 0.021 | 0.026 | 0.011 | 0.015 | 0.014 | 0.014 |
| **By Age** |  |  |  |  |  |  |
| 17 - 20 | 0.017 | 0.032 | 0.008 | 0.017 | 0.009 | 0.008 |
| 20 - 29 | 0.023 | 0.024 | 0.012 | 0.014 | 0.014 | 0.014 |
| 30 - 39 | 0.023 | 0.031 | 0.012 | 0.017 | 0.016 | 0.016 |
| 40 - 49 | 0.020 | 0.022 | 0.011 | 0.014 | 0.014 | 0.011 |
| 50 - 59 | 0.019 | 0.021 | 0.010 | 0.013 | 0.017 | 0.015 |
| 60 - 84 | 0.016 | 0.026 | 0.012 | 0.018 | 0.015 | 0.015 |
| **By Race** |  |  |  |  |  |  |
| Black | 0.021 | 0.026 | 0.011 | 0.015 | 0.014 | 0.013 |
| Other | 0.009 | 0.029 | 0.009 | 0.019 | 0.004 | 0.019 |
| White | 0.026 | 0.028 | 0.013 | 0.015 | 0.019 | 0.021 |
| **By Floor** |  |  |  |  |  |  |
| Floor 1 | 0.023 | 0.033 | 0.007 | 0.016 | 0.028 | 0.017 |
| Floor 2 | 0.049 | 0.071 | 0.034 | 0.048 | 0.040 | 0.043 |
| Floor 3 | 0.029 | 0.015 | 0.013 | 0.011 | 0.013 | 0.010 |
| Floor 4 | 0.008 | 0.016 | 0.004 | 0.005 | 0.008 | 0.011 |
| Floor 5 | 0.015 | 0.017 | 0.003 | 0.008 | 0.011 | 0.007 |
| Floor 6 | 0.009 | 0.010 | 0.006 | 0.006 | 0.002 | 0.002 |
| Floor 7 | 0.016 | 0.015 | 0.011 | 0.007 | 0.001 | 0.002 |

<sup>1</sup> The number of times a cell-level location spell ends without its corresponding activity spells also ending, divided by the average active network size, divided again by the number of days in the timeframe of interest (for scaling purposes). The result is an estimate of the number of cell changes per person per day.
<sup>2</sup> The number of times a block-level location spell ends without its corresponding activity spells also ending, divided by the average active network size, divided again by the number of days in the timeframe of interest (for scaling purposes). The result is an estimate of the number of block changes per person per day.
<sup>3</sup> The number of known releases (i.e., the number of spells of activity that are not right-censored), divided by the average active network size, divided again by the number of days in our timeframe of interest (for scaling purposes). The result is an estimate of the number of daily releases per resident.

## DISCUSSION

In this study, we provided one of the first empirical descriptions of contact networks within a carceral environment during an ongoing outbreak of a respiratory infectious disease with airborne transmission. Our estimates over two COVID-19 timespans (during the peak of the Omicron wave, and 4 months after it) for network degree, mixing, and dissolution by age group, race/ethnicity, and floor number can be useful for understanding the transmission potential of respiratory infectious diseases like SARS-CoV-2 and the real-world responses to them. Our model summary statistics can also be used for parameterizing network-based mathematical models that simulate the impact of biomedical and non-pharmaceutical interventions, such as quarantining, jail depopulation, and vaccination.

Overall, we found that the network characteristics of jail residents were likely to be highly conducive to the spread of SARS-CoV-2 and other infectious diseases. This was reflected in several network characteristics. We observed a very high cross-sectional network degree at both the cell and block levels, reflecting the fact that most residents shared a cell with another resident and that cell blocks aggregated many cells. The cumulative distribution of the cell-level network was right-skewed due to some residents sleeping in common quarters within the cell block due to cell capacity constraints. These conditions create ideal contact situations for the spread of respiratory diseases, both due to the high network degree at the block level and the intensity of contacts at the cell level. Prior research has noted high numbers of contacts within carceral environments, but not to the level of network specificity as our current study (6).

The rapid growth in the temporal networks would likely facilitate the spread of disease (28). This growth was the result of the turnover in the cell-level networks, block-level networks, and jail population overall. Jails, in contrast to prisons, are inherently comprised of residents with higher rates of population turnover due to their role as shorter-term facilities to hold persons awaiting trial and those with lesser criminal charges. This, in addition to the need for resident movement within the jail for general population control, creates environments ideal for the spread of infectious diseases like SARS-CoV-2 as infectious residents may transitions into the jail and across cell-blocks within a short time period (23). Extensions of our dynamic network degree measures have been developed, including a forward reachable path measure that tracks the evolving links between network members as network edges form and dissolve over time (29).

Another important contribution of our study was to evaluate changes in network structure during a SARS-CoV-2 outbreak and after it had passed. This comparison suggested that the jail administration lessened some of the risk of morbidity for the residents, although more could have been done. We found that the mean degree was 40% lower among older residents during the Omicron peak, and 20% lower overall. This suggests that some quarantining was implemented, lowering risk of SARS-CoV-2 acquisition generally and COVID disease for those older residents who were more likely to fare worse with infection. Accounting for network dissolution, dynamic network degree growth (Figure 5) was also lower during Omicron compared to after it at both the cell-level and block-level networks. However, the size of the dynamic network degree during the Omicron peak suggests high potential for disease spread. Finally, during both timespans, there was strongly assortative age mixing, but segregating residents by age did not seem to be purposefully undertaken. Our analysis suggests that age segregation may have further limited the spread of disease to older residents.

### Limitations

This analysis had some limitations. First, we only report on the contact networks between residents, and not for staff (e.g., correctional officers) also in the jail. This was due to that our data source, cell roster data, was only available for residents and not for others. Our analysis therefore underestimates the true contact network for residents, particularly as staff enter and exit the jail each day. Second, we assume that spatial co-location represents a contact, and therefore a network “edge” for our summary statistics. Spatial co-location within a jail cell, even for a short period of time, would constitute an infection-probable contact for SARS-CoV-2 and other infectious diseases (i.e., an infection event would occur with some non-zero probability given this co-location). However, the transmission potential for block-level edges is less clear. This is partly because all members of a cell block rarely congregate together, and in many cases, this is prohibited due to resident control. However, because block-level contacts may represent a non-zero transmission probability contact type, we elected to represent them separately within these analyses. Future modeling studies could explore on cell-level versus block-level transmission under different assumptions of transmission probabilities. Finally, there were certain days in the analysis time frame for which jail rosters were unavailable. This could generate interval censoring, which would impact dissolution rate calculations. For this reason, we focused on two time periods of 11 days with consecutive rosters for many analyses.

### Conclusions

Infectious diseases like SARS-CoV-2 have had a major impact on the health of persons in jail and prisons, both due to the dense contact network conditions within these settings and the underlying risk factors of persons before incarceration. Limited empirical data was available to characterize the patterns of contacts within a large and diverse jail setting. Our study provided some of the first rigorous network statistics, both to describe these network patterns generally and also to compare how networks changed under conditions of a major COVID outbreak period. We hope that this information is useful in understanding contact patterns to support future research and public health activities within these settings.

## Data Availability

All data produced in the present study are available upon reasonable request to the authors.

## ACKNOWLEDGEMENTS

The research team wishes to thank the Fulton County Sheriff and jail staff, especially Records Manager Ms. Kielah Yancey, for their invaluable assistance.

